# Effects of Ultrasound Cycloplasty following Failed Surgery for Glaucoma: 12-months Results From a Prospective Clinical Study

**DOI:** 10.1101/2025.03.05.25323432

**Authors:** Liu Li, Ying-Jie Li, Ling Hong, Yong-Bo Wang, Xuan Zhu

**Author notes:** Correspondence: Ying-Jie Li, Department of Ophthalmology, Nanchang First Hospital, Nanchang 330038, China.

## Abstract

**Objectives:** This prospective study aims to evaluate the outcomes and safety of Ultrasound Cycloplasty(UCP) in controlling intraocular pressure in patients with prior failed glaucoma surgeries.

**Methods:** A total of 20 eyes from 19 patients who underwent UCP following failed glaucoma surgery between September 2020 to September 2022, were included. All patients were followed for over 12 months. Intraocular pressure, ocular hypotensive medications, and best-corrected visual acuity were recorded and compared after surgery. Complete success was defined as intraocular pressure within the range of 6-21mmHg after treatment, without the use of anti-glaucoma agents. Qualified success was defined the same as complete success but allows the presence of anti-glaucoma agents.

**Results:** At 12 months follow-up, intraocular pressure decreased from 45.10±10.74 mmHg to 20.80±6.05 mmHg, representing a reduction of 53.88% (P < 0.01). The rates of complete successes and qualified successes were 20.00% (4 of 20 eyes) and 65.00% (13 of 20 eyes) 12 months after treatment respectively.

**Conclusion:** UCP is a safe and effective surgical approach for eyes with prior failed glaucoma surgeries.

## Introduction

Glaucoma is a common, irreversible blinding eye disease characterized by optic nerve damage and progressive visual field defects [1]. The most effective treatment to mitigate vision loss is controlling intraocular pressure(IOP) [2].

However, there are still many patients who cannot achieve target IOP deemed safe for the eye. Trabeculectomy, glaucoma drainage device implantation, and cyclodestructive procedure are three commonly used treatment methods. But there may be several limitations. Repeat trabeculectomy may result in choroidal effusion, shallow or flat anterior chamber and wound leak [3].Implantation of glaucoma drainage device may brings a risk of diplopia, corneal edema and tube erosions [4].

Additionally, cyclodestructive procedures can lead to significant risks, including inflammation, low intraocular pressure, vision loss, and even phthisis bulbi due to non-selective destruction of target tissues and unpredictable dose-response relationships [5,6]. Therefore, there is an urgent need to explore more effective and safer treatment options to address the issue of persistent IOP following failed glaucoma surgeries.

Ultrasound cycloplasty (UCP) provides a new approach for the cyclodestructive procedure. UCP utilizes the highly selective effect of HIFU on the ciliary body, achieving an automated computer-assisted ciliary body treatment program. The main mechanisms of focused ultrasound in the treatment of glaucoma are as follows: 1) coagulation of ciliary epithelial cells and reduction of aqueous humor secretion [7]; and 2) reorganization of scleral tissue in the treatment area leads to separation of sclera and ciliary body, thus increasing outflow of aqueous humor from the suprachoroidal space [8]. Its application and repetition do not increase the risk of complications [9]. The purpose of this study was to explore safety and efficacy of UCP following failed surgery for glaucoma at 12 month.

## Materials and methods

### Patients

This is a prospective single-center study. Patients who underwent UCP between September 2020 to September 2022 following failed glaucoma surgery were consecutively included. All patients signed a written informed consent form, and this study has been approved by the Ethics Committee of Nanchang First Hospital.

Inclusion criteria was as followings: (1) Age 18-80 years, no gender limitation. (2) Single or multiple prior failed glaucoma surgeries followed by UCP. (3) Baseline IOP≥21mmHg, regardless of anti-glaucoma medication use. (4) Minimum 12 months of follow-up. Exclusion criteria was as followings:(1) History of ocular infections and uveitis; (2) Normal tension glaucoma; (3) Thin sclera or ocular tumor; (4) Patients with severe systemic diseases previously or currently who cannot tolerate UCP treatment.

### UCP Procedure

All surgical procedures were performed by a same experienced ophthalmologist (Ying-Jie Li). Patients received treatment with an appropriate size of probe which was selected according to the distance of horizontal ciliary process of UBM and white to white of IOL Master before treatment by machine (EyeOP1, Eye Tech Care, France). The skin around the surgical eye was disinfected routinely before treatment, the conjunctival sac was rinsed with povidone iodine and physiological saline, and treatment was performed under retrobulbar anesthesia with lidocaine. An appropriate range of the UCP sector was chosen according to patient’s preoperative IOP, visual acuity, and degree of optic nerve atrophy [10]. 3:00 o’clock and 9:00 o’clock positions were avoided to protect from the anterior ciliary artery and posterior ciliary nerve branches damage, as shown in the figure 1.

**Figure 1.**
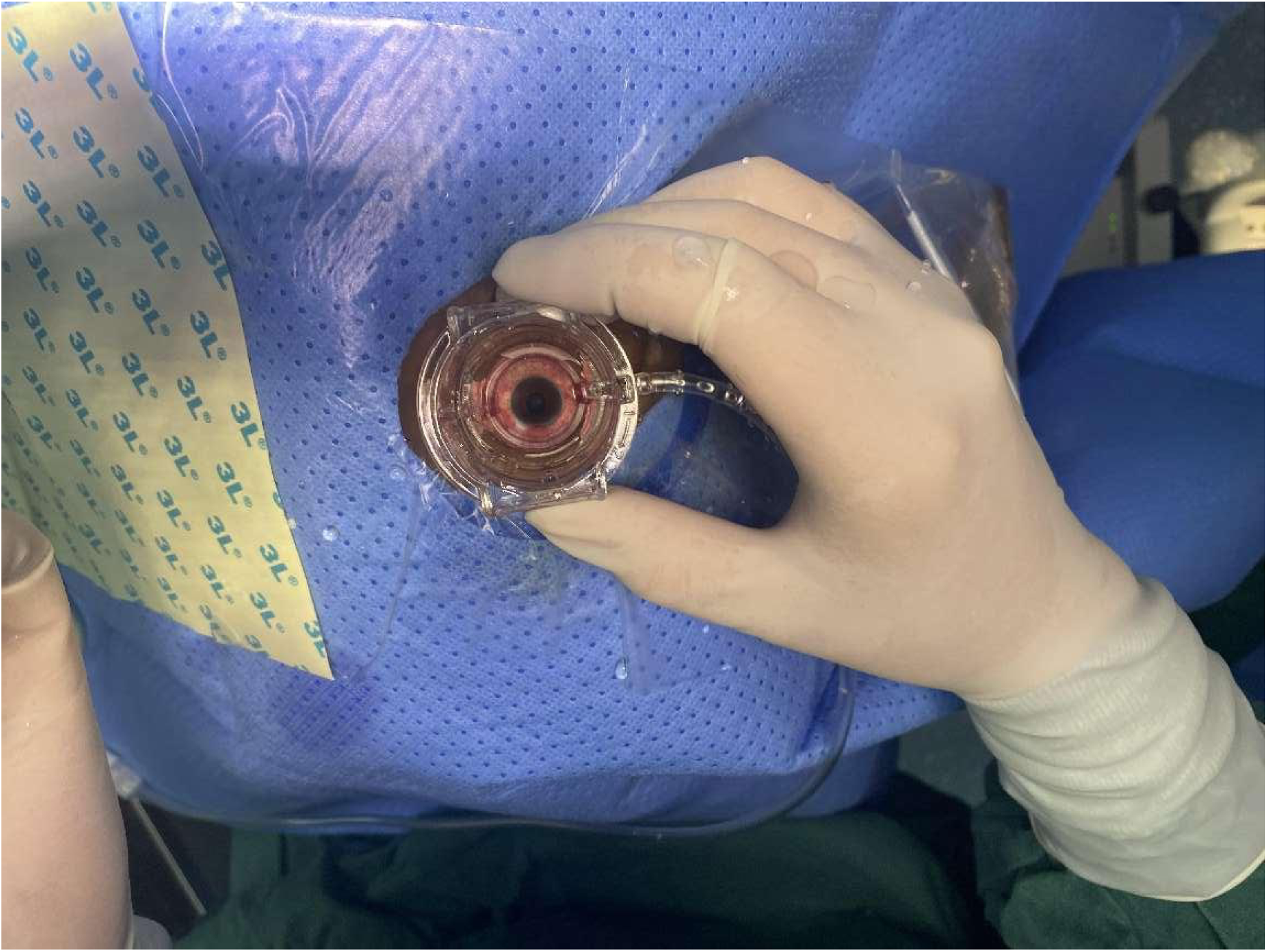
The surgeon was positioned on the patient’s head, and performing treatment on the right eye.

**Figure 2.**
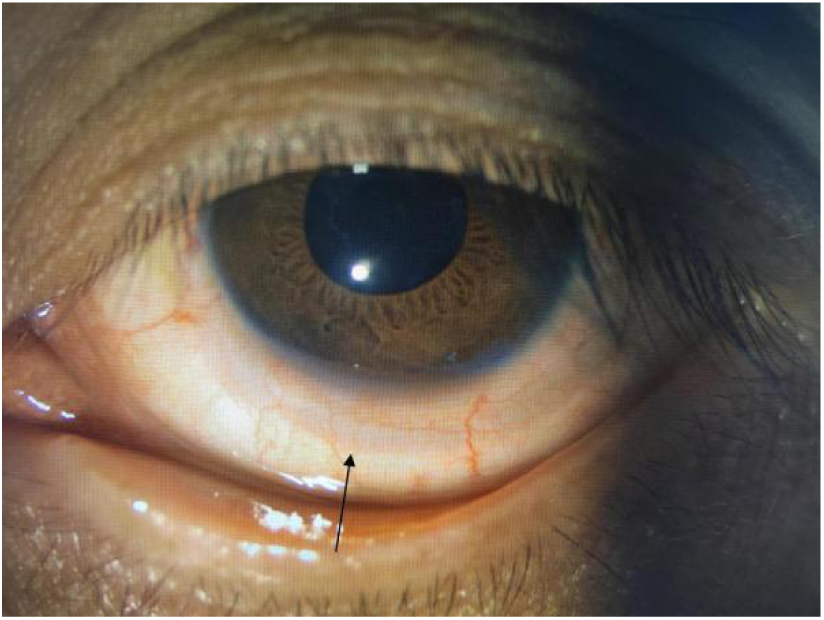
Scleral imprint observed 12 months after UCP treatment. Black arrow shows scleral imprint.

### Followed-up

After UCP treatment, praprofen eye drops and tobramycin dexamethasone eye drops (4 times/day) were administered to the eyes, and the number of ocular hypotensive medications was adjusted according to the patient’s intraocular pressure situation. After treatment the preoperative ocular hypotensive medications were maintained if the intraocular pressure was higher than 21mmHg. The use of prostaglandin drugs was discontinued first, followed by beta blockers if the intraocular pressure was between 10-21mmHg. And all the ocular hypotensive medications were stoped if the intraocular pressure was lower than 10mmHg. Patients were followed up for 12 months after treatment. Intraocular pressure, the number of ocular hypotensive medications, best corrected visual acuity (LogMAR) before and 1 day, 1 week, 1, 3,6 and 12 months after treatment were recorded. The Snellen BCVA data were converted to LogMar, which represents the angle in the smallest resolution unit. The visual acuity for counting fingers, hand movements, light perception (LP), and no light perception (NLP) were recorded as 2.0, 2.4, 3.0, and 3.5 respectively [11]. And the complications during treatment and after treatment were observed.

### Surgical Success

Complete success: Treatment was considered to be complete successful when IOP after treatment was between 6 and 21mmHg without anti-glaucoma agent or eventual retreatment.

Qualified success: The criteria was same as complete success but allow the use of ocular hypotensive medications.

### Statistical Analysis

SPSS 25.0 software (IBM, USA) was used for statistical analysis. Measurement data was represented as mean ± standard deviation. Paired t-tests were conducted to compare postoperative data with preoperative data. *P*<0.05 was considered to be statistically significant.

## Results

### Demographic and Characteristics of the Patients

As detailed in Table 1, this study included 9 males (10 eyes) and 11 females (11 eyes), aged 24-79 years, with an average age of (57.95 ± 14.19) years, had undergone an average of 1.60±0.60 glaucoma surgeries before treatment. Based on previous medical records and gonioscopic examination, 9 eyes of primary angle-closure glaucoma (45.00%), 5 eyes of secondary open-angle glaucoma (25.00%, with 1 intravitreal silicone oil injection, 2 glucocorticoid-induced, 2 angle recession), 3 eyes of primary open-angle glaucoma (15.00%), and 3 eyes of secondary angle-closure glaucoma (15.00%, with 3 neovascular) were selected. There were 4 eyes (20.00%) with moderate glaucoma, 11 eyes (55.00%) with severe glaucoma and 5 eyes (25.00%) with absolute glaucoma.

**TABLE 1.**
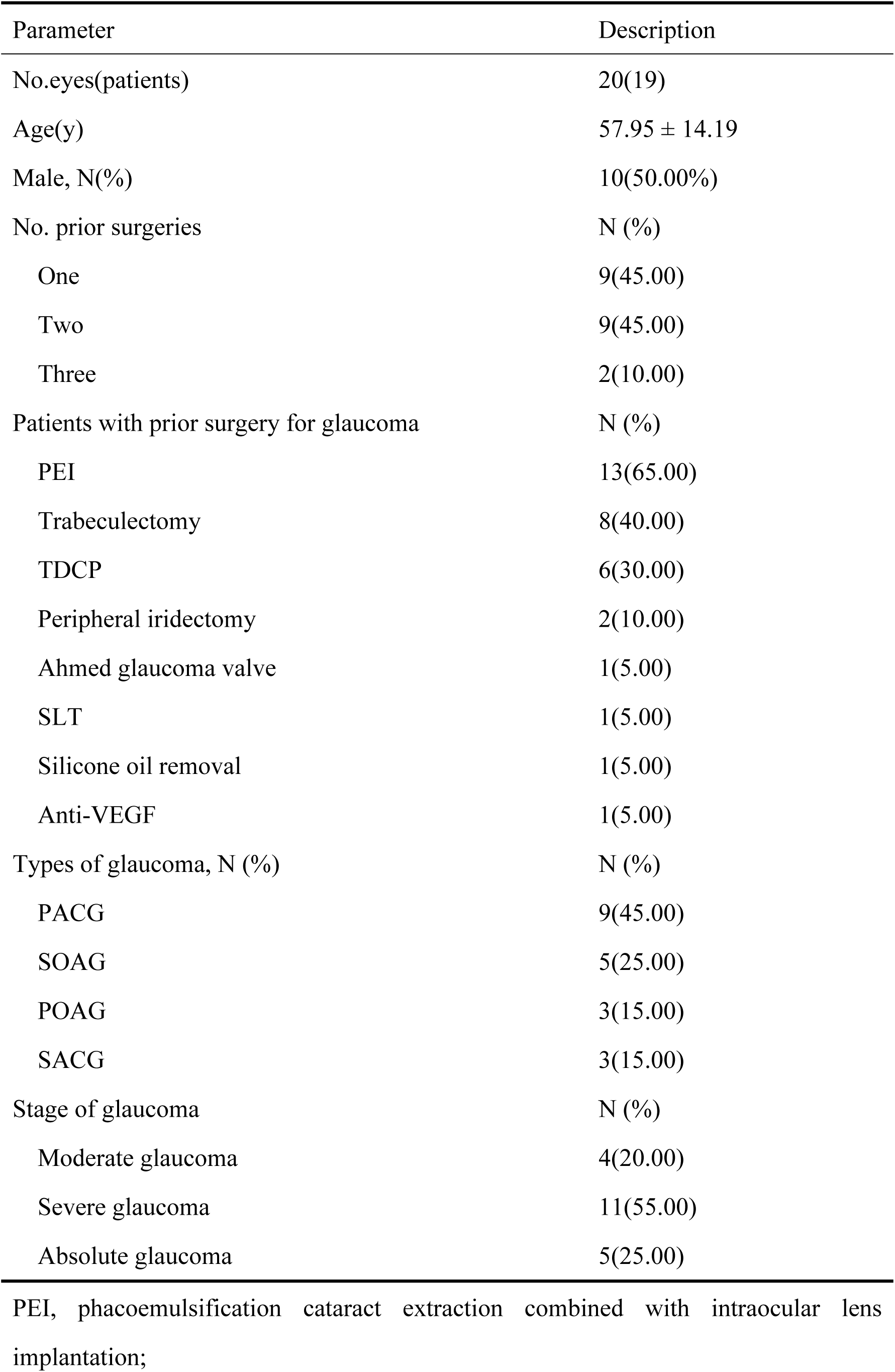
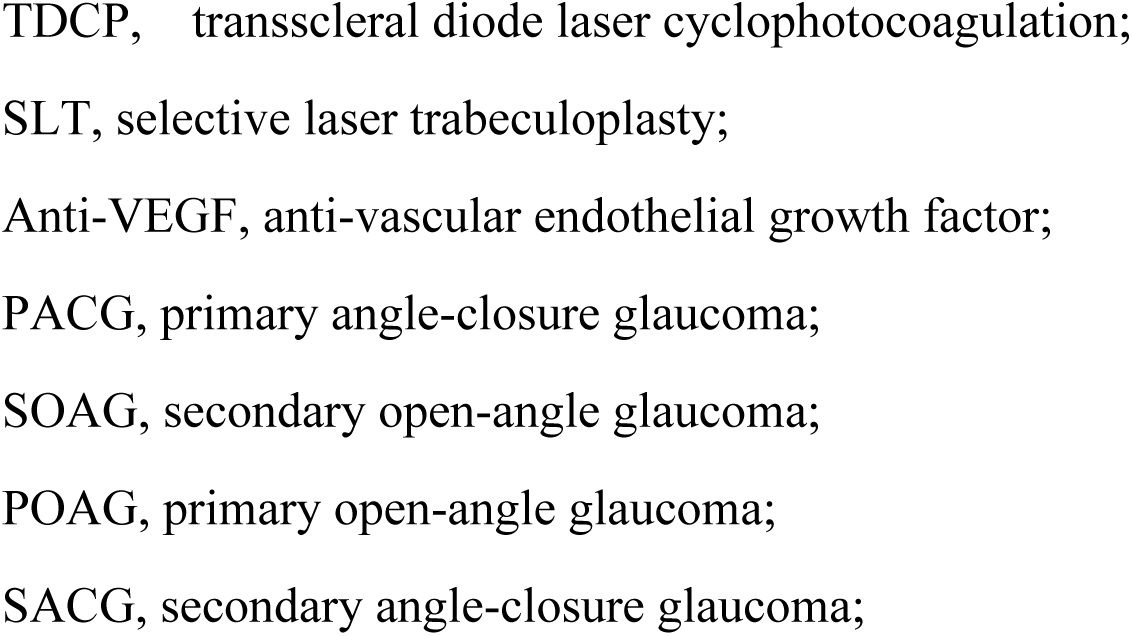
Baseline Demographics and Characteristics of the Study Participants.

| Parameter | Description |
| --- | --- |
| No.eyes(patients) | 20(19) |
| Age(y) | 57.95 ± 14.19 |
| Male, N(%) | 10(50.00%) |
| No. prior surgeries | N (%) |
| One | 9(45.00) |
| Two | 9(45.00) |
| Three | 2(10.00) |
| Patients with prior surgery for glaucoma | N (%) |
| PEI | 13(65.00) |
| Trabeculectomy | 8(40.00) |
| TDCP | 6(30.00) |
| Peripheral iridectomy | 2(10.00) |
| Ahmed glaucoma valve | 1(5.00) |
| SLT | 1(5.00) |
| Silicone oil removal | 1(5.00) |
| Anti-VEGF | 1(5.00) |
| Types of glaucoma, N (%) | N (%) |
| PACG | 9(45.00) |
| SOAG | 5(25.00) |
| POAG | 3(15.00) |
| SACG | 3(15.00) |
| Stage of glaucoma | N (%) |
| Moderate glaucoma | 4(20.00) |
| Severe glaucoma | 11(55.00) |
| Absolute glaucoma | 5(25.00) |

PEI, phacoemulsification cataract extraction combined with intraocular lens implantation;
TDCP, transscleral diode laser cyclophotocoagulation;
SLT, selective laser trabeculoplasty;
Anti-VEGF, anti-vascular endothelial growth factor;
PACG, primary angle-closure glaucoma;
SOAG, secondary open-angle glaucoma;
POAG, primary open-angle glaucoma;
SACG, secondary angle-closure glaucoma;

### Changes in IOP, Number of Anti-Glaucoma Medications, and BCVA

The preoperative intraocular pressure of patients was 22-57 (mean 45.10±10.74) mmHg. IOP decreased from 45.10±10.74 mmHg at baseline to 20.80±6.05 mmHg at month 12(P˂0.05). The number of ocular hypotensive medications decreased from 2.95±1.23 at baseline to 2.30±1.53 at month 12. (Table 2)

**Table 2.** Changes of IOP, Number of Medications of the Study Participants.

| Time | No. eyes | IOP(mmHg) | P(IOP) | No. medications | P(medications) |
| --- | --- | --- | --- | --- | --- |
| Pre- | 20 | 45.10±10.74 |  | 2.95±1.23 |  |
| D 1 | 20 | 25.75±9.24 | <0.01 | 0.90±1.37 | <0.01 |
| Wk 1 | 20 | 18.90±6.70 | <0.01 | 0.95±1.00 | <0.01 |
| Mo 1 | 20 | 19.40±7.43 | <0.01 | 1.20±1.11 | <0.01 |
| Mo 3 | 20 | 20.50±6.62 | <0.01 | 1.45±1.23 | <0.01 |
| Mo 6 | 20 | 20.00±7.58 | <0.01 | 2.10±1.48 | 0.023 |
| Mo 12 | 20 | 20.80±6.05 | <0.01 | 2.30±1.53 | 0.085 |

Data were presented as mean (standard deviation) .
IOP, intraocular pressure.

BCVA showed no significant change during the entire follow-up process compared to preoperative levels (P> 0.05). (Table 3)

**Table 3.** Changes of BCVA of the Study Participants.

| Time | No. eyes | BCVA (Logmar) | P | t |
| --- | --- | --- | --- | --- |
| Pre- | 20 | 1.89±1.26 |  |  |
| D 1 | 20 | 1.81±1.31 | 0.617 | 0.508 |
| Wk 1 | 20 | 1.82±1.29 | 0.642 | 0.473 |
| Mo 1 | 20 | 1.80±1.28 | 0.602 | 0.530 |
| Mo 3 | 20 | 1.75±1.26 | 0.061 | 1.990 |
| Mo 6 | 20 | 1.77±1.22 | 0.204 | 1.316 |
| Mo 12 | 20 | 1.86±1.28 | 0.739 | 0.339 |

Data were presented as mean (standard deviation) .
IOP, best corrected visual acuity.

### Success Rate and Complications

The rates of complete successes and qualified successes were 20.00% (4 of 20 eyes) and 65.00% (13 of 20 eyes) 12 months after treatment respectively.

There was no serious complication during the study period. Four eyes experienced mild to moderate pain during the treatment, yet they still cooperated to complete the treatment. The postoperative complications were relatively mild, and most of them disappeared within one month spontaneously or after medication. While scleral imprint lasted a long time and were still observed 12 months after UCP treatment (Table 4).

**Table 4.** Intra-operative and postoperative complications.

| Description | Number of eyes (%) |
| --- | --- |
| Intraoperative |  |
| Pain | 4(20.00) |
| Post-operative |  |
| Scleral imprint | 11(55.00) |
| Conjunctival hyperemia | 10(50.00) |
| Anterior chamber inflammation | 1(4.35) |
| Hyphema | 1(4.35) |

## Discussion

UCP achieved success rates of 65.00% at month 12. It was notably that there was still a significant decrease in IOP(a reduction of 53.88%), with no serious complications observed. A 20% reduction in IOP significantly reduces the risk of future visual field loss [12],making UCP a feasible surgical option for glaucoma patients. These findings are consistent with previous research on the efficacy and safety of UCP following failed glaucoma surgery [9,13,14].

UCP, as an initial surgery, had shown good results in the treatment of glaucoma. Almobarak FA [15] et al studied the outcomes of UCP for the first surgery for glaucoma, reporting mean IOP decreased from 23.16±6.4mmHg on 3.27 ± 0.9 ocular hypotensive medications preoperatively to 16.57±6.0mmHg on 1.86±1.4 ocular hypotensive medications at the 12-months visit. The efficacy of UCP also sustains for a long time. A study on UCP revealed that after three years, the reduction of IOP still maintained 10.6 mmHg with a success rate of 55% [16]. Similarly, Wang T [17] et al studied the efficacy of UCP for 36 patients with refractory glaucoma, reporting a mean reduction rate of 43.05% 6 months after UCP. These findings suggest that UCP has an effect comparable to that of the initial surgery in maintaining IOP following failed surgery for glaucoma.

Ruixue W [18] et al compared UCP and ECP for glaucoma with a 12 months follow-up, they found that UCP and ECP have comparable intraocular pressure lowering effects after surgery, but UCP has fewer postoperative complications. Another research revealed that UCP and cyclocryotherapy both showed significant efficacy in reducing intraocular pressure. However, UCP treatment was safer, with fewer postoperative complications and adverse reactions [19]. Micro-pulse transscleral cyclophotocoagulation(MP-TSCPC) may be a safe non-invasive technique for lowering IOP in a variety of glaucoma patients, including those who do not experience a satisfactory decrease in IOP after prior surgery. However, the success rate of the initial MP-TSCPC was below 25%, while the success rate for repeat surgeries does not exceed 45% [20]. Compared to other minimally invasive glaucoma surgeries, UCP is an effective and safe option.

The rates of complete and qualified successes were 20.00% (4 of 20 eyes) and 65.00% (13 of 20 eyes) 12 month after treatment respectively which were lower than Deb [21]. There may be several reasons for this: First, the subjects included in this study were all refractory glaucoma, which may explain the poor treatment outcome. Second, some patients included in this study had no light perception before treatment, so they refused to take anti-glaucoma agent after treatment, leading to high intraocular pressure and thus considered a failure. Third, the criteria for success were not entirely the same.

Scleral imprint was a relatively rare complication for UCP treatment in some studies [22], while it was common in this study. Some researchers suggested that scleral imprint was a sign of scleral thinning [21]. But in this study, all patients with scleral imprints showed no sign of scleral thinning, such as staphyloma until the end of follow-up. An underwent UBM examination on the scleral imprint found that sclera was not obviously thinner but interlayer structure was sparse [23]. Rodolfo Mastropasqua found formation of new (or the enlargement of preexisting) intrascleral hyporeflective spaces after UCP treatment through anterior segment optical coherence tomography [8]. We suggest there may be a histological rearrangement, which may explain the gray appearance found under slit lamp. It deserves our attention and long-term observation.

The limitations of this study include: 1) The sample size was relatively small; 2) We did not compare the efficacy with other ciliary body destructive surgeries or glaucoma surgeries; 3) Heterogeneity in glaucoma types and baseline IOP levels. Further research is in progress to investigate IOP control with long term follow-up.

## Conclusion

UCP is successful in achieving IOP reduction with excellent safety for the treatment of patients who had prior failed surgery for glaucoma.

## Author’ Contribution

All author contributed to the conception of this study. Liu Li contributed to data analysis and wrote the manuscript. All surgical procedures were performed by Ying-Jie Li. Ling Hong collected the data. Yong-Bo Wang elaborated the figures. Xuan Zhu contributed to the interpretation. All authors read and approved the final manuscript.

## Declaration

## Consent for publication

Not Applicable.

## Ethical approval

All experimental protocols were approved by Ethics Committee of Nanchang First Hospital. Informed consent was obtained from all subjects.

## Availability of data and materials

The data that support the findings of this study are available from the corresponding author upon reasonable request.

## Data Availability

Data cannot be shared publicly because of patient's Privacy. Data are available from the Nanchang First Hospital (contact via Liu Li) for researchers who meet the criteria for access to confidential data.

## References

1. Jonas JB, Aung T, Bourne RR, Bron AM, Ritch R, Panda-Jonas S. Glaucoma. Lancet. 2017;390 (10108):2183–2193.

2. Liang YB, Jiang JH, Wang NL. A review of epidemiological investigation and research on glaucoma in China. Chin J Ophthalmol, 2019, 55 (8): 634–640.

3. Gedde SJ, Herndon LW, Brandt JD, Budenz DL, Feuer WJ, Schiffman JC; Tube Versus Trabeculectomy Study Group. Postoperative complications in the Tube Versus Trabeculectomy (TVT) study during five years of follow-up. Am J Ophthalmol. 2012 May;153(5):804–814.e1. doi: 10.1016/j.ajo.2011.10.024. Epub 2012 Jan 14. PMID: 22244522; PMCID: PMC3653167.

4. Rockwood EJ. The Ahmed Baerveldt Comparison (ABC) Study: Long-Term Results, Successes, Failures, and Complications. Am J Ophthalmol. 2016 Mar;163:xii-xiv. doi: 10.1016/j.ajo.2015.12.031. Epub 2016 Jan 29. PMID: 26830056.

5. Nassiri N, Syeda S, Tokko H, Thipparthi M, Cohen MI, Kim C, Al-Timimi FR, Tannir JR, Goyal A, Juzych MS, Hughes BA, Wilson MR. Three-year outcomes of trabeculectomy and Ahmed valve implant in patients with prior failed filtering surgeries. Int Ophthalmol. 2020 Dec;40(12):3377–3391. doi: 10.1007/s10792-020-01525-y. Epub 2020 Aug 10. PMID: 32776301.

6. Benson MT, Nelson ME. Cyclocryotherapy: a review of cases over a 10-year period. Br J Ophthalmol. 1990 Feb;74(2):103–5. doi: 10.1136/bjo.74.2.103. PMID: 2310722; PMCID: PMC1042001.

7. Cao H, Xu Z, Long H, et al. Trans-catheter arterial chemoembolization in combination with high-intensity focused ultrasound for unresectable hepatocellular carcinoma: a systematic review and meta-analysis of the Chinese literature. Ultrasound Med Biol 2011; 3 (7): 1009–1016.

8. Mastopasqua R, Agnifili L, Fasanella V, et al. Uveo-scleral outflow pathways after ultrasonic cyclocoagulation in refractory glaucoma: an anterior segment optical coherence tomography and in vivo confocal study. Br J Ophthalmol 2016; 100 (12): 1668–1675.

9. Morais Sarmento T, Figueiredo R, Garrido J, Passos I, Rebelo AL, Candeias A. Ultrasonic circular cyclocoagulation prospective safety and effectiveness study. Int Ophthalmol. 2021;41 (9):3047–3055.

10. Fan FF, Ge X, Liu DD, Xu TY, Wang RX, Chen XY, Li SY. Comparison of the efficacy and safety of ultrasonic cycloplasty vs valve implantation and anti-VEGF for the treatment of fundus disease-related neovascular glaucoma. Int J Ophthalmol. 2023 Jun 18;16(6):897–903. doi: 10.18240/ijo.2023.06.10. PMID: 37332547; PMCID: PMC10250935.

11. Luo Q, Xue W, Wang Y, Chen B, Wang S, Dong Y, Ru Y, Ge L. Ultrasound Cycloplasty in Chinese Glaucoma Patients: Results of a 6-Month Prospective Clinical Study. Ophthalmic Res. 2022;65(4):466–473. doi: 10.1159/000515013. Epub 2021 Feb 4. PMID: 33540418.

12. Chauhan BC, Mikelberg FS, Artes PH, Balazsi AG, LeBlanc RP, Lesk MR, Nicolela MT, Trope GE; Canadian Glaucoma Study Group. Canadian Glaucoma Study: 3. Impact of risk factors and intraocular pressure reduction on the rates of visual field change. Arch Ophthalmol. 2010 Oct;128(10):1249-55. doi: 10.1001/archophthalmol.2010.196. Epub 2010 Aug 9. Erratum in: Arch Ophthalmol. 2010 Dec;128(12):1633. PMID: 20696979.

13. Aptel F, Tadjine M, Rouland JF. Efficacy and Safety of Repeated Ultrasound Cycloplasty Procedures in Patients With Early or Delayed Failure After a First Procedure. J Glaucoma. 2020 Jan;29(1):24–30. doi: 10.1097/IJG.0000000000001400. PMID: 31842139.

14. Yu Q, Liang Y, Ji F, Yuan Z. Comparison of ultrasound cycloplasty and transscleral cyclophotocoagulation for refractory glaucoma in Chinese population. BMC Ophthalmol. 2020 Sep 29;20(1):387. doi: 10.1186/s12886-020-01655-y. PMID: 32993561; PMCID: PMC7525941.

15. Almobarak FA, Alrubean A, Alsarhani WK, Aljenaidel A, Osman EA. Two-Year Outcomes of Ultrasound Cyclo Plasty as a First Procedure in Glaucoma. Semin Ophthalmol. 2023 Jul;38(5):482–489. doi: 10.1080/08820538.2023.2170715. Epub 2023 Feb 10. PMID: 36762779.

16. Rouland JF, Aptel F. Efficacy and Safety of Ultrasound Cycloplasty for Refractory Glaucoma: A 3-Year Study. J Glaucoma. 2021 May 1;30(5):428–435. doi: 10.1097/IJG.0000000000001796. PMID: 33900251.

17. Wang T, Wang R, Su Y, Li N. Ultrasound cyclo plasty for the management of refractory glaucoma in chinese patients: a before-after study. Int Ophthalmol. 2021 Feb;41(2):549–558. doi: 10.1007/s10792-020-01606-y. Epub 2020 Oct 18. PMID: 33070270.

18. Ruixue W, Wenjun D, Le J, Fangfang F, Ning L, Xiaoya C, Suyan L. A comparative study of ultrasound cycloplasty and endoscopic cyclophotocoagulation in the treatment of secondary glaucoma. Sci Rep. 2023 Dec 27;13(1):23073. doi: 10.1038/s41598-023-50157-6. PMID: 38155225; PMCID: PMC10754948.

19. Ruixue W, Tao W, Ning L. A Comparative Study between Ultrasound Cycloplasty and Cyclocryotherapy for the Treatment of Neovascular Glaucoma. J Ophthalmol. 2020 Jan 22;2020:4016536. doi: 10.1155/2020/4016536. PMID: 32047661; PMCID: PMC7001673.

20. Hooshmand S, Voss J, Hirabayashi M, McDaniel L, An J. Outcomes of initial and repeat micro-pulse transscleral cyclophotocoagulation in adult glaucoma patients. Ther Adv Ophthalmol. 2022 Feb 14;14:25158414211064433. doi: 10.1177/25158414211064433. PMID: 35187401; PMCID: PMC8852201.

21. Deb-Joardar N, Reddy KP. Application of high intensity focused ultrasound for treatment of open-angle glaucoma in Indian patients. Indian J Ophthalmol 2018; 66(4): 517–523.

22. De Gregorio, A., Pedrotti, E., Stevan, G. et al. Safety and efficacy of multiple cyclocoagulation of ciliary bodies by high-intensity focused ultrasound in patients with glaucoma. Graefes Arch Clin Exp Ophthalmol 2017;255, 2429–2435.

23. Zhou LF, Hu Die, Lan Jie, et al. Efficacy and safety of single Ultrasound Cyclo-Plasty to treat refractory glaucoma: Results at 1 year. Eur J Ophthalmol. 2022 Jan;32(1):268–274. doi: 10.1177/1120672120973605. Epub 2020 Nov 22. PMID: 33225725.

